# Impact of smartphone-enabled home urinary albumin-to-creatinine ratio testing on albuminuria screening and management

**DOI:** 10.1101/2025.06.11.25329440

**Authors:** Waleed Zafar, Yirui Hu, Lauren Brubaker, Nathaniel Harshaw, Jamie Green, Alexander R Chang

**Author notes:** Corresponding author: Dr. Alexander R. Chang, MD, MS, Center for Kidney Health Research, Departments of Nephrology and Population Health Sciences, Geisinger Medical Center, 100 North Academy Avenue, Danville, PA 17822.

## Abstract

**Objective:** We investigated the impact of smartphone-enabled home urinary albumin creatinine ratio (UACR) testing on albuminuria screening and management in a prospective propensity score-matched cohort study.

**Research design and methods:** We randomly selected 4000 adult individuals (50% with hypertension but no diabetes; 50% with diabetes) receiving primary care at a large, regional health system to receive Minuteful Kidney, an FDA-cleared, smartphone-enabled home UACR test kit. A total of 3998 individuals in the intervention program were matched 1:1 to 3998 in the control group receiving usual care, using propensity score matching including sociodemographics and comorbidities. The primary outcome was completion of UACR test (either lab-based or home testing) within 100 days of program start.

**Results:** Overall, completion of any UACR test was higher in the intervention program than the control group for the hypertension subgroup (53% vs. 13%) and the diabetes subgroup (53% vs. 30%). Increase in UACR testing associated with Minuteful Kidney was consistent across age, sex, race, and ethnicity subgroups though lower in non-users of the patient portal. Minuteful Kidney UACR results were abnormal (≥30 mg/g) in 38% (330/872) in the hypertension subgroup and 45% (306/682) in the diabetes subgroup. Among Minuteful Kidney-tested individuals, those with abnormal UACR were more likely to have follow-up primary care and nephrology visits and new prescriptions of renin-angiotensin-aldosterone system inhibitors than those with normal UACR.

**Conclusion:** A convenient smartphone-enabled home albuminuria test is effective in increasing albuminuria screening among high-risk individuals.

**Twitter summary:** Home urine tests using smartphones boosted kidney screening rates 2.5x vs. usual care. Early detection can result in better follow-up and treatment to prevent heart and kidney problems. #kidneyhealth

Why did we undertake this study?

- We undertook this study to see if we could improve early detection of kidney disease through at-home albuminuria screening.

What is the specific question(s) we wanted to answer?

- We wanted to know whether a smartphone-enabled home urine test could increase screening rates.

What did we find?

- Home testing significantly increased test completion (53.1% vs. 21.2%), and there were some early suggestions of improved diagnosis and follow-up care.

What are the implications of our findings?

- Home testing for albuminuria can enhance early detection of kidney disease, which could lead to earlier identification, management and potentially long-term outcomes.

## INTRODUCTION

Chronic kidney disease (CKD) is a major public health problem in the United States affecting more than 1 in 7 U.S adults (1). CKD is associated with an increased incidence of end stage kidney disease (ESKD) and cardiovascular events (2,3). The individual and societal burdens of CKD are high (4), with an estimated annual Medicare treatment cost for CKD of $87 billion and for ESKD of an additional $37.3 billion (5). Presence of albuminuria plays a crucial role in effective risk stratification of individuals with hypertension (HTN) and diabetes mellitus (DM). Early detection of albuminuria can enable use of indicated therapeutic interventions such as angiotensin converting enzyme inhibitors (ACEis) or angiotensin receptor blockers (ARBs), and sodium glucose cotransporter-2 inhibitors (SGLT2is) to reduce the risk of progression to ESKD and cardiovascular complications thus underscoring the significance of timely diagnosis and management.

The Kidney Disease Improving Global Outcomes (KDIGO) 2024 guidelines recommend albuminuria testing among individuals at high risk for CKD including those with HTN, DM and cardiovascular disease (CVD) (6). KDIGO and American Diabetes Association (ADA) guidelines recommend at least annual urine albumin/creatinine ratio (UACR) testing for individuals with DM (7,8) and CKD (individuals with CKD may need more frequent testing) (6). The European Society of Cardiology / European Society of Hypertension recommends UACR testing at least once for those with HTN (9) while the American Heart Association (AHA) 2017 HTN guidelines suggests UACR testing as an ‘optional’ test to provide information about target organ damage (10). However, an updated hypertension guideline is under development, and the AHA also includes albuminuria as an additional test to further enhance risk prediction of cardiovascular disease in the AHA Predicting Risk of CVD EVENTs (PREVENT) calculator (11).

However, despite the clear clinical and economic effectiveness of albuminuria screening in at-risk populations, adherence to testing remains very low. Only 40% of individuals with diabetes and 10% of those with hypertension are estimated to complete testing at the recommended frequency (2,3). Alarmingly, a substantial number of these patients have not undergone UACR measurements in the past year, indicating a notable gap in their care.

Several barriers to albuminuria screening have been described (12). One possible barrier to screening for albuminuria in primary care settings is that urine sample collection at the primary care provider’s office may not be feasible or convenient. Home based testing can thus improve screening. Initial evaluation of smartphone-based home urine protein screening among pregnant women showed the test to be feasible (13). In an earlier randomized controlled trial, we have shown that smartphone-based urine protein testing is feasible and results in higher rates of screening compared to usual care among individuals with hypertension (14). Use of home testing kits for proteinuria screening among pediatric and young adult population using the vendor Healthy.io has also been found to be associated with high satisfaction among patients and their caregivers (15). Studies using the same vendor and kit in the Netherlands have also shown high user satisfaction (16). The sensitivity of smartphone-enabled semi-quantitative UACR home test has been found to be comparable to laboratory-based UACR testing in an analysis of 555 urine samples (sensitivity 96.2%, specificity 84.2% and negative predictive value of 98.6% and positive predictive value of 66.8%) (17). In this study, we examined the impact of a prospective propensity score-matched cohort study evaluating the impact of a smartphone-enabled home UACR test as a population health strategy to improve albuminuria screening. We hypothesized that among at-risk individuals (those with HTN and/or DM) due for albuminuria screening, an enhanced strategy using Minuteful Kidney (“intervention program”) would result in higher adherence to albuminuria screening compared to a propensity score-matched cohort receiving usual care (“control program”).

## METHODS

This study included adult individuals receiving primary care at Geisinger, a large integrated, mostly rural health system in central and northeast Pennsylvania providing care to approximately 1.2 million people. This study was funded by Boehringer Ingelheim and was approved by the Geisinger Institutional Review Board (IRB 2023-1788).

### Study population

The study population were adults 18-85 years of age receiving primary care at Geisinger who were diagnosed with either HTN or DM, who had not undergone UACR testing in the prior 12 months and who had a valid US address and phone number on file. We randomly selected 4000 adults (50% with HTN but no DM and 50% with DM +/-HTN) to receive Minuteful Kidney, an FDA-cleared, smartphone-enabled home urine albuminuria test kit. We excluded individuals with end-stage kidney disease (ESKD) International Classification of Diseases (ICD-10) codes, estimated glomerular filtration rate (eGFR) < 15 ml/min/1.73m^2^, and individuals receiving palliative care, or in long-term nursing care per health maintenance modifiers or problem list codes. To address potential confounding due to non-random treatment assignment, we performed 1:1 nearest-neighbor propensity score matching without replacement and identified an equal number of controls stratified by sub-cohorts of HTN only and DM (+/-HTN). Matching was done for baseline covariates including age, sex, race, ethnicity, body mass index (BMI), HgbA1c (for DM patients), HTN diagnosis (for DM patients), insurance type, Charlson Comorbidity Index, atherosclerotic cardiovascular disease, congestive heart failure (CHF), and estimated glomerular filtration rate (eGFR).

### Intervention

Participants who were randomly selected to receive Minuteful Kidney were informed about this research study by the test vendor (Healthy.io) via an introduction email, text messages, and letter, elaborating about the study purpose and design. Healthy.io’s engagement service (including calls, text messages, letters, emails) were used to assist participants to complete the test as required. Patients had the option to opt out of the communication channels as well as the study at any point without any consequences. The Minuteful Kidney package included a welcome letter with information about the service, why they were receiving it, guidance on completing the test, information regarding the Minuteful Kidney smartphone application and contact numbers for troubleshooting and to opt out of the research study.

The smartphone application uses interactive video, text, and sound instruction to guide users through the testing procedure. Software with an augmented reality layer on the smartphone camera guides users while taking the scan, presents the UACR test results immediately to the patient, and recommends next steps to guide patients in arranging follow-up with their PCPs for abnormal test results. The testing kit consists of a standard urinalysis dipstick, a custom designed urine cup and a color-board, which enables accurate analysis in different lighting environments. To conduct the test, patients open the application, follow directions provided on-screen, collect urine in the provided container, dip the urinalysis dipstick, place the dipstick on the color board, and then scan the dipstick and color board using the application and the camera on their mobile device.

The Minuteful Kidney UACR test results were uploaded to the Geisinger electronic health record (Epic) by e-fax through Health Insurance Portability and Accountability Act (HIPAA)-compliant routine clinical processes at Geisinger. The test results were carbon copied to patients’ primary care providers. The research team verified incoming UACR test results weekly with Healthy.io and Geisinger staff to ensure accuracy and completeness of data transmission. As this study was designed to determine the impact of the Minuteful Kidney service, there were no specific actions taken by the study team or additional support staff to aid with CKD management. However, the nephrologist investigators (ARC, JG) answered questions from providers promptly with suggestions to follow KDIGO CKD guidelines (e.g. use of ACEi/ARB, SGLT2i, blood pressure control <130/80 mmHg for albuminuria).

### Outcomes

The primary outcome was completion of any UACR test (either Minuteful Kidney or laboratory based UACR test) within 100 days from the start of patient outreach by the test vendor (i.e. send out of test kits that commenced 02/04/2024). Secondary outcomes included completion of kidney health evaluation (i.e. at least one eGFR and one UACR within 12 months), and actions taken by the participants’ care team within 12 months of the start of the patient outreach including appointments with primary care providers, nephrology appointments, nephrology e-consults, new ICD diagnoses of CKD or proteinuria, and new prescriptions of cardiorenal-protective medications including ACEi/ARBs, SGLT2 inhibitors, nonsteroidal mineralocorticoid receptor antagonists (MRAs) and steroidal MRAs. Among individuals who completed Minuteful Kidney tests, we also examined retesting of abnormal Minuteful Kidney UACR tests by regular laboratory UACR testing within 12 months, and patient engagement and satisfaction surveys that were administered after completion of their tests. Manual chart review of individuals with abnormal Minuteful Kidney tests were done at 4 months after the start of the patient outreach (starting from 06/04/2024) by a medical student (NH) to gain insights regarding EHR-documented contact between patients and providers.

### Statistical Analyses

#### Sample size determination

With a sample size of 4000 individuals receiving the intervention and similar number of controls, we estimated that we would have 80% power at a 0.05 significance level to detect a 2.9% change in the albuminuria screening rates from a baseline value of 10% (control) to 12.9% (intervention), corresponding to an odds ratio of 1.33.

#### Propensity score matching

We matched individuals who received home albuminuria testing to controls receiving usual care using propensity score with 1:1 ratio and nearest-neighbor matching for age groups (18 to <50, 50 to <60, 60 to <70, 70-85), sex, race, ethnicity, insurance type, Charlson Comorbidity Index, eGFR categories (using quantile-based binning), atherosclerotic cardiovascular disease, and CHF. Continuous variables with skewed distributions (such as age and eGFR) were transformed into categorical bins using clinical cut-offs or quantile bins. Individuals with missing values will be excluded from the matching. A caliper of 0.1 standard deviation was applied in the matching algorithm. Covariate balance between matched groups was evaluated using standardized mean difference (SMD), with SMD < 0.1 indicating acceptable balance.

We compared the outcomes between the two matched groups using McNemar’s test. Baseline characteristics were compared using t-tests or Mann-Whitney tests for continuous variables and Chi-square or Fisher’s exact tests for categorical variables. A p-value <0.05 was considered statistically significant. Statistical analyses were conducted in RStudio (version).

## RESULTS

A total of 3998 individuals who were enrolled in the Minuteful Kidney program (“intervention program”) were matched to 3998 individuals who received usual care (“control program”) with no significant differences between groups (Table 1). Mean age was 61 years (standard deviation 13.6 years), 51% were male, 93% were white, 4% were of Hispanic or Latino ethnicity, 50% had diabetes, 76% had hypertension, 18% had atherosclerotic CVD; and 12% had CHF. Mean BMI was 33 kg/m^2^.

**Table 1:** Baseline demographic characteristics for Intervention program and Control program.

|  | Intervention Program | Control Program | p-value |
| --- | --- | --- | --- |
| n | 3998 | 3998 |  |
| Age in years; Mean (SD) | 61.17 (13.6) | 61.06 (13.6) | 0.723 |
| Male; n (%) | 2031 (50.8) | 2021 (50.6) | 0.84 |
| White; n (%) | 3718 (93.0) | 3723 (93.1) |  |
| Hispanic / Latino; n (%) | 177 (4.4) | 165 (4.1) |  |
| Not Hispanic or Latino; n (%) | 3723 (93.1) | 3737 (93.5) |  |
| BMI; Mean (SD) | 33.18 (8.2) | 33.33 (8.3) | 0.411 |
| Type of Insurance |  |  | 0.99 |
| Geisinger Health Plan | 764 (19.1) | 756 (18.9) |  |
| Geisinger Health Plan Gold (Medicare Advantage) | 718 (18.0) | 706 (17.7) |  |
| Medicaid | 196 (4.9) | 199 (5.0) |  |
| Medicare | 753 (18.8) | 758 (19.0) |  |
| Medicare Advantage | 318 (8.0) | 329 (8.2) |  |
| Military | 21 (0.5) | 24 (0.6) |  |
| Other commercial plans | 1228 (30.7) | 1226 (30.7) |  |
| Chronic disease score; mean (SD) | 2.04 (1.91) | 2.05 (1.92) | 0.838 |
| Diabetes mellitus ICD diagnosis; n (%) | 1999 (50.0) | 1999 (50.0) | 1 |
| HTN ICD diagnosis; n (%) | 3023 (75.6) | 3023 (75.6) | 1 |
| ASCVD ICD diagnosis; n (%) | 707 (17.7) | 720 (18.0) | 0.726 |
| CHF ICD diagnosis; n (%) | 459 (11.5) | 462 (11.6) | 0.944 |
| Any CKD; n (%) | 753 (18.8) | 779 (19.5) | 0.481 |
| 2021 eGFR; mean (SD) | 84.08 (20.3) | 84.04 (20.7) | 0.921 |
| Baseline ACEi or ARB use; n (%) | 2362 (59.1) | 2309 (57.7) | 0.233 |
| Baseline SGLT2 inhibitor use; n (%) | 423 (10.6) | 365 (9.1) | 0.032 |
| Baseline nonsteroidal MRA use; n (%) | 150 (3.8) | 156 (3.6) | 0.772 |
| Baseline steroidal MRA use; n (%) | 0 | 0 |  |
| BMI: Body mass index; eGFR: estimated glomerular filtration rate; HTN: Hypertension; ASCVD: Atherosclerotic cardiovascular disease; CHF; Congestive Heart Failure; |  |  |  |

### UACR Testing Outcomes

Overall, UACR test completion rate across at-risk untested patients was 53.1% among those in the intervention program and 21.2% among those in the control program (p<0.001). Improvement in UACR testing associated with the intervention program was higher in the HTN-only cohort (40.9% absolute difference between intervention and control groups) than in the DM +/-HTN cohort (22.7% absolute difference between intervention and control programs) (Table 2). The higher UACR testing rates associated with the intervention program were fairly consistent across age groups, sex, race and ethnicity (Table 3). However, the intervention program had less of an impact among patients who did not use the Epic MyChart^®^ patient portal (absolute between-arm difference 12%) compared to those who were patient portal users (absolute between-arm difference 37%). At one year from the start of the study (Feb 05, 2025), 68.0% in the intervention program vs. 46.7% in the control program had at least one UACR test done and 62.5% in the intervention program vs. 45.0% in the control program had both an eGFR and a UACR test done.

**Table 2:**
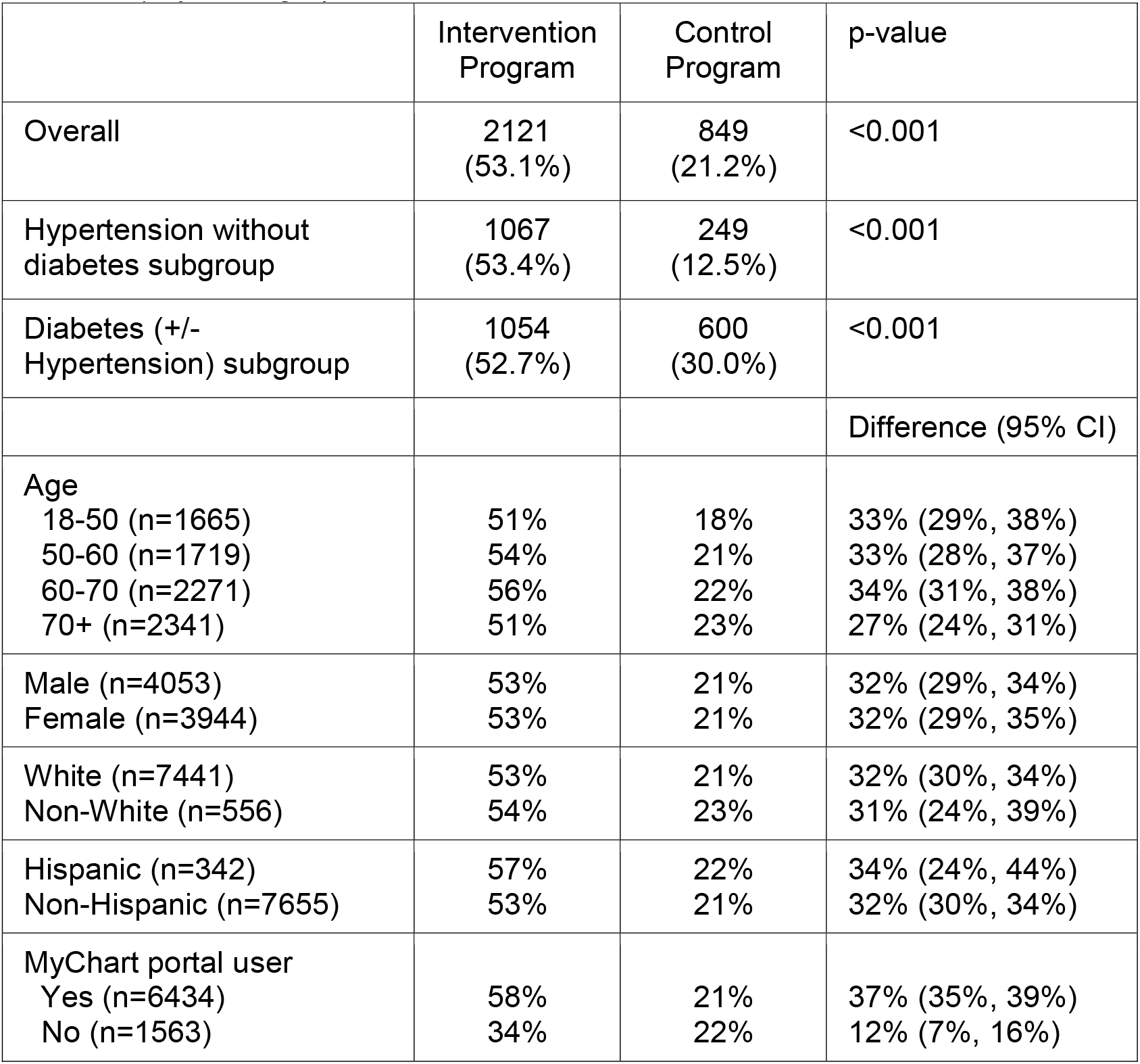
Improvement in ACR screening at 100 days from the study initiation (primary outcome) by demographics.

**Table 3:** Secondary care process outcomes among Intervention Program and Control Program.

|  | Overall<br>N=7997 | Intervention<br>Program<br>N=3998 | Control<br>Program<br>N=3999 | p value |
| --- | --- | --- | --- | --- |
| Primary care, completed (%) | 7108 (88.9) | 3572 (89.3) | 3536 (88.4) | 0.202 |
| Nephrology, completed (%) | 244 (3.1) | 133 (3.3) | 111 (2.8) | 0.172 |
| E-consult with nephrology, Completed (%) | 21 (0.3) | 9 (0.2) | 12 (0.3) | 0.663 |
| ACEi or ARBs, new Rx; n (%) | 447 (5.6) | 209 (5.2) | 238 (6.0) | 0.174 |
| SGLT2i, new Rx; n (%) | 322 (4.0) | 159 (4.0) | 163 (4.1) | 0.866 |
| MRA, new Rx; n (%) | 110 (1.4) | 51 (1.3) | 59 (1.5) | 0.502 |
| nsMRA, new Rx; n (%) | 3 (0.0) | 1 (0.0) | 2 (0.1) | 1 |
| CKD or proteinuria ICD*, new diagnosis; n (%) | 303 (3.8) | 188 (4.7) | 115 (2.9) | <0.001 |
| Proteinuria ICD, new diagnosis; n (%) | 138 (1.7) | 106 (2.7) | 32 (0.8) | <0.001 |
| CKD ICD, new diagnosis; n (%) | 173 (2.2) | 88 (2.2) | 85 (2.1) | 0.877 |
| ACEi Angiotensin converting enzyme inhibitors; ARB Angiotensin receptor blockers; CKD Chronic kidney disease; MRA Mineralocorticoid receptor antagonists; nsMRA non-steroidal Mineralocorticoid receptor antagonists; SGLT2i Sodium-Glucose Cotransporter-2 inhibitors; |  |  |  |  |

\* Defined by ICD codes
Proteinuria - Proteinuria R80.\*
Any CKD diagnosis - CKD stage 1 N18.1, CKD stage 2 N18.2, CKD stage 3 N18.3, CKD stage 4 N18.4, CKD stage 5 N18.5, End-stage kidney disease N18.6, Kidney transplant status Z94.0, Unspecified CKD stage N18.9, Diabetic CKD E08.22, E09.22, E10.22, E11.22, E13.22, Hypertensive CKD I12.\*, I13.\*

Among those in the intervention program who were tested by Minuteful Kidney, there were roughly similar rates of albuminuria detected in the hypertension subgroup (UACR 30-300 mg/g: 281/872 [32.2%]; >300 mg/g: 49/872 [5.6%]) and the diabetes subgroup (UACR 30-300 mg/g: 235/682 [34.5%]; >300 mg/g: 71/682 [10.4%]) (Supplemental Table 1,2). Among individuals who received Minuteful Kidney testing, those with moderately increased A2 albuminuria (UACR 30-300 mg/g) and severely increased A3 albuminuria (UACR >300 mg/g) results were more likely (64.3% and 66.7% respectively) to have had a quantitative follow-up UACR test compared to individuals with normal Minuteful Kidney test result (40.2%). Specifically, the corresponding odds ratios (OR) of a follow-up UACR were OR 2.68 (95% CI: 2.15, 3.36) for those with A2 albuminuria group, and OR 2.97 (95% CI: 2.00, 4.48) for those with A3 albuminuria, as compared to individuals with normal Minuteful Kidney test results (UACR <30 mg/g).

### Actions taken by the participants’ care team

Overall, there were more new CKD or proteinuria ICD diagnoses in the intervention program than in the control program (4.7% vs. 2.9%; p<0.001). There were no significant differences in new prescriptions of ACEi or ARB, SGLT2, or mineralocorticoid medications between the two programs (p>0.05 for comparisons) (Table 4).

**Table 4:**
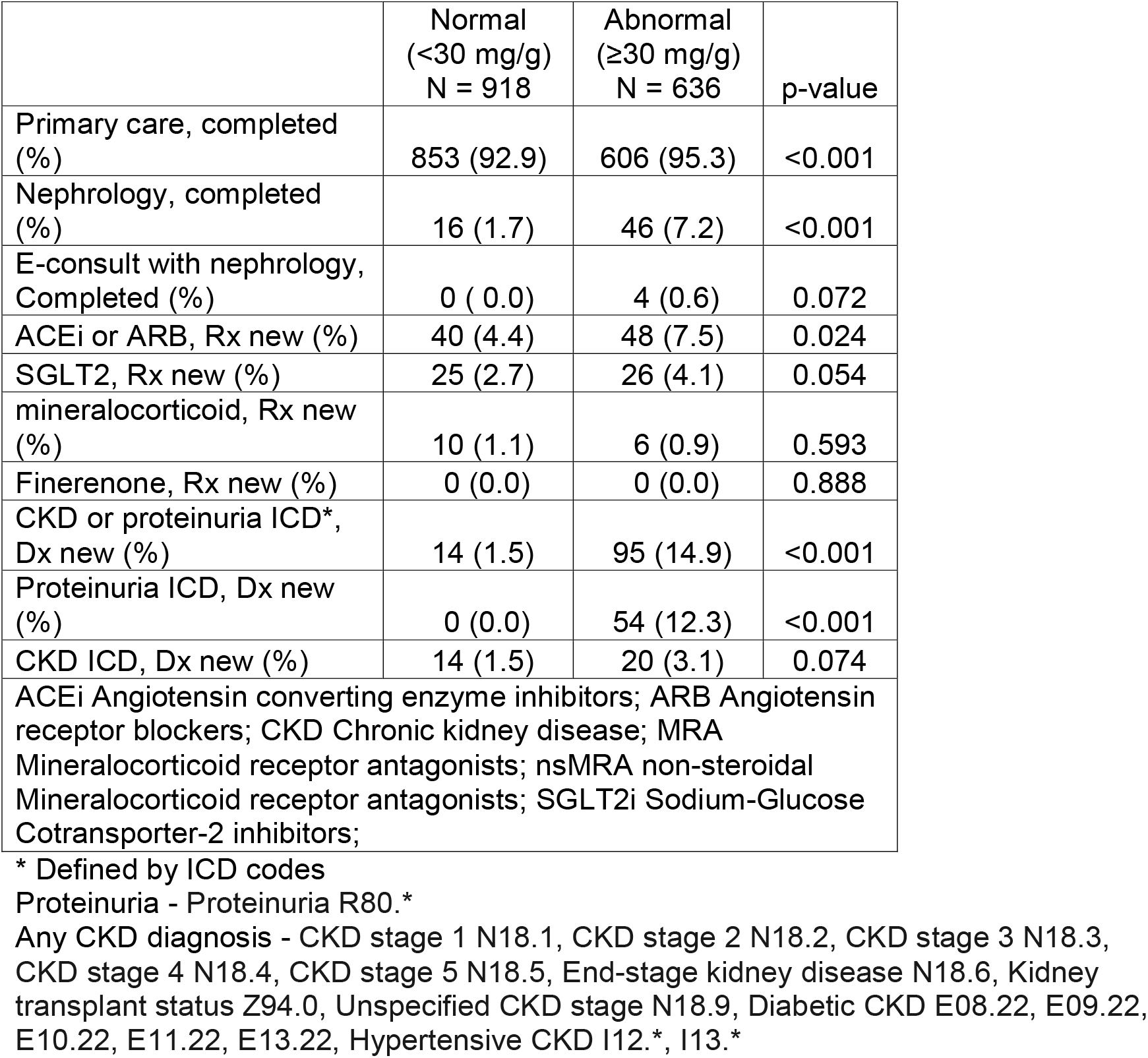
Secondary outcomes among those who completed Minuteful Kidney home ACR test stratified by test result at 1 year from initiation of the study.

| | Normal<br>($<30$ mg/g)<br>N = 918 | Abnormal<br>($\geq 30$ mg/g)<br>N = 636 | p-value |
| --- | --- | --- | --- |
| Primary care, completed (%) | 853 (92.9) | 606 (95.3) | $<0.001$ |
| Nephrology, completed (%) | 16 (1.7) | 46 (7.2) | $<0.001$ |
| E-consult with nephrology, Completed (%) | 0 (0.0) | 4 (0.6) | 0.072 |
| ACEi or ARB, Rx new (%) | 40 (4.4) | 48 (7.5) | 0.024 |
| SGLT2, Rx new (%) | 25 (2.7) | 26 (4.1) | 0.054 |
| mineralocorticoid, Rx new (%) | 10 (1.1) | 6 (0.9) | 0.593 |
| Finerenone, Rx new (%) | 0 (0.0) | 0 (0.0) | 0.888 |
| CKD or proteinuria ICD*, Dx new (%) | 14 (1.5) | 95 (14.9) | $<0.001$ |
| Proteinuria ICD, Dx new (%) | 0 (0.0) | 54 (12.3) | $<0.001$ |
| CKD ICD, Dx new (%) | 14 (1.5) | 20 (3.1) | 0.074 |
| ACEi Angiotensin converting enzyme inhibitors; ARB Angiotensin receptor blockers; CKD Chronic kidney disease; MRA Mineralocorticoid receptor antagonists; nsMRA non-steroidal Mineralocorticoid receptor antagonists; SGLT2i Sodium-Glucose Cotransporter-2 inhibitors; |  |  |  |

\* Defined by ICD codes
Proteinuria - Proteinuria R80.\*
Any CKD diagnosis - CKD stage 1 N18.1, CKD stage 2 N18.2, CKD stage 3 N18.3, CKD stage 4 N18.4, CKD stage 5 N18.5, End-stage kidney disease N18.6, Kidney transplant status Z94.0, Unspecified CKD stage N18.9, Diabetic CKD E08.22, E09.22, E10.22, E11.22, E13.22, Hypertensive CKD I12.\*, I13.\*

However, among intervention program individuals who completed testing with Minuteful Kidney, those with an abnormal result (UACR≥30 mg/g) were significantly more likely to have a new CKD or proteinuria ICD diagnosis (14.9% vs. 1.5%; p<0.001), complete a follow-up primary care visit (95.3% vs. 92.9%; p<0.001), complete a follow-up nephrology visit (7.2% vs. 1.7%; p<0.001), start a new ACEi or ARB (7.5% vs. 4.4%; p=0.02), and start a new SGLT2 inhibitor (4.1% vs. 2.7%; p=0.05), compared to those with a normal Minuteful Kidney result (UACR <30 mg/g) (Table 5). Findings were similar in hypertension subgroup and diabetes subgroup analyses (Supplemental Tables 1,2).

### EHR-documentation of patient-provider communication after abnormal Minuteful Kidney tests

After 16 weeks from initiation of the study, we sought to characterize patient-provider communications after the receipt of abnormal Minuteful Kidney test results (Supplemental Figure 1). Among the 636 individuals with positive Minuteful Kidney tests, 322 (50.6%) had test-related communication between patients and their providers documented in the electronic health record. The vast majority (266 out of 322; 83%) were patient initiated, and the remainder were provider initiated. Among those that were initiated by the patient, 40.6% were via MyChart portal, 37% were mentioned during an appointment, and 22.5% were over the phone.

### Minuteful Kidney-captured Patient Surveys

After completion of Minuteful Kidney, 890 patients (22.3%) completed a survey. Satisfaction was high among individuals in the Minuteful Kidney program with a 53+ Net Promoter Score. The vast majority (94%) rated the test as easy or very easy to complete, and 71% stated preference for at home testing (Supplemental Table 3). No significant adverse events were reported. However, 6 (0.2%) patients reached out to the team to ask about the authenticity of the intervention and the Minuteful Kidney test kit with most being satisfied after discussion with the team. One patient expressed having serious concerns about data privacy and data sharing.

## DISCUSSION

In this prospective quasi-randomized propensity-score matched cohort study of adult primary care patients with hypertension or diabetes who had not received albuminuria testing in the past year, we found that a smartphone-based home albuminuria testing strategy was highly successful in improving albuminuria screening. Individuals in the HTN-only (without DM) cohort saw a greater improvement (53.4% vs. 12.5% in control) than those in the DM+/-HTN cohort (52.7% vs. 30.0% in control). Increase in the UACR testing associated with Minuteful Kidney was consistent across age groups, sex, race and ethnicity though the impact was lower among patients who did not use the patient portal compared to those who were active patient portal users.

Following completion of testing, about half of the patients had documented interactions with their providers in the EHR with 83% of these interactions initiated by the patient. These suggest the testing may have been successful in activating patients in their own care. Among those with albuminuria detected, there tended to be increased use of new ACEi or ARB prescription and new SGLT2 inhibitor prescription, compared to those who had normal albuminuria Minuteful Kidney test results. We also found that among the intervention program, there was increased proteinuria diagnoses but no significant change in CKD diagnoses. This is consistent with earlier work showing that CKD remains underdiagnosed using ICD diagnosis codes (18,19) in part because of physicians’ lack of awareness that proteinuria with eGFR>60 constitutes CKD I/II (20).

A Healthcare Effectiveness Data and Information Set (HEDIS) measure, Kidney Health Evaluation for Patients with Diabetes (KED) was adopted in 2020, promoting the completion of both an eGFR and an ACR test result annually (21). While we found that use of the Minuteful Kidney test significantly improved annual completion of an eGFR and an ACR test, it should be noted that the current HEDIS measure does not include semiquantitative ACR as a qualifying test for the measure. KDIGO 2024 CKD guidelines do include semiquantitative ACR as a testing option, acknowledging that it may be less accurate than a quantitative test but still would be better than missing a measure (6). We believe that use of the test in a multimodal population health approach to capture patients with missing ACR test results could be useful. The sensitivity of smartphone-enabled semi-quantitative UACR home test has been found to be comparable to laboratory-based UACR testing in an analysis of 555 urine samples (sensitivity 96.2%, specificity 84.2% and negative predictive value of 98.6% and positive predictive value of 66.8%) with 85.6% of samples accurately classified into KDIGO albuminuria categories (17).

During the course of our study, we learned several important lessons related to implementation. Despite multiple announcements to the primary care group about the intervention, we still had a handful of patients reaching out to inquire about the validity of the program, and one patient who was upset about privacy with the communication from the vendor despite reassurances that the study was IRB-approved and followed study protocol policies appropriately with a waiver of consent. Patient concerns about data privacy and data sharing have been noted previously in a similar study (22). We believe that future efforts will need to ensure better integration of such programs with the EHR, or consider physician opt-in to the study, or more integrated communications between the vendor and the health system. For example, in this pilot study there was no associated lab order in the electronic health record, and we did not use the patient portal to communicate with patients.

Timely diagnosis of albuminuria can identify individuals in need of more intensive therapy as treatments such as ACEis, ARBs, and SGLT2is are recommended to reduce cardiorenal risk. In addition, the American Heart Association (AHA)’s Predicting Risk of CVD EVENTs (PREVENT) calculator can incorporate albuminuria to enhance CVD risk prediction (11). By improving risk stratification of CVD and CKD risks, this type of population health intervention has the potential to have downstream benefits on cardiovascular and kidney health though larger studies with longer follow-up would be needed to demonstrate this. A major strength of our study was the real-world population health approach deployment, and excellent fidelity in delivering the intervention and capturing results back into the EHR. Use of an experienced vendor’s patient engagement processes was essential as well as an engaged population health team. While not technically a randomized controlled study, we randomly assigned patients to receive the intervention and prospectively created a well-matched control group. A limitation was that the population was mostly white, mirroring the studied population in central and northeast Pennsylvania. Further studies would be helpful to generalize findings to other settings and additional work is needed to improve management once albuminuria is detected.

### Conclusions

A population health approach using smartphone-enabled home albuminuria testing was effective in increasing albuminuria screening among individuals with hypertension or diabetes with potential benefits in improved CKD recognition and treatment of detected albuminuria.

## Data Availability

All data produced in the present study are available upon reasonable request to the corresponding author

## Acknowledgements

Funding and Assistance: This study was funded by Boehringer-Ingelheim. Boehringer Ingelheim was given the opportunity to review the manuscript for medical and scientific accuracy, as well as intellectual property considerations. However, the authors have full jurisdiction over all aspects of study design, conduct, analysis, and publication.

## Author contributions

WZ writing initial draft (lead); YH performed statistical analyses; LB conceptualization (supporting), project administration; NH data collection; JG conceptualization (supporting); ARC conceptualization (lead), writing initial draft (supporting), project administration, supervision. All authors reviewed and edited the manuscript. ARC is the guarantor of this work and, as such, had full access to all the data in the study and takes responsibility for the integrity of the data and the accuracy of the data analysis.

## Prior presentation

Some of these data were presented at the American Heart Association Hypertension Scientific Sessions 2024 on 9/6/24, and an oral abstract presentation presenting data in this manuscript is planned for the American Diabetes Association 85^th^ Scientific Sessions June 20^th^, 2025.

