## Supplemental Tables for "Impact of smartphone-enabled home urinary albumin-to-creatinine ratio testing on albuminuria screening and management"

**Online-only Supplemental Material**

Supplemental Figure 1. Patient provider interactions among Minuteful Kidney program participants

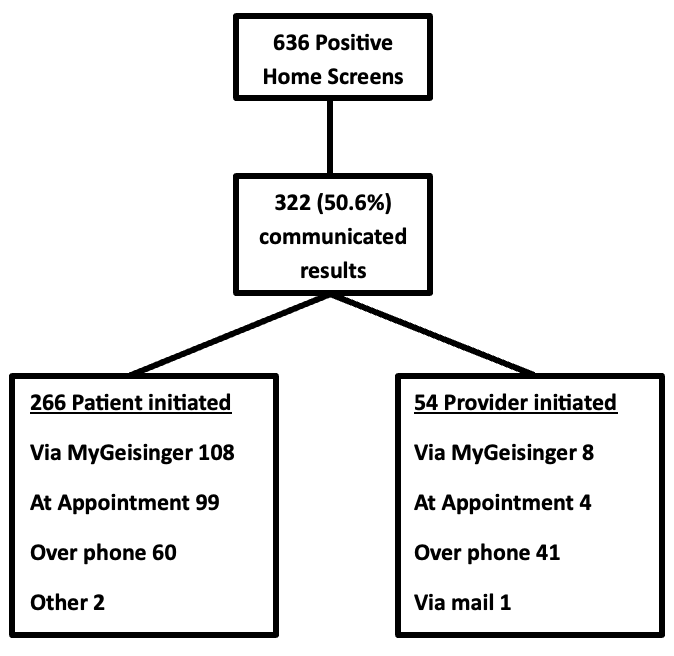

Supplemental Table 1: Secondary outcomes among those in the Hypertension (no diabetes) subgroup who completed Minuteful Kidney home ACR test stratified by test result at 1 year from initiation of the study

|  | Normal  (<30 mg/g)  N = 542 | Abnormal (30-300 mg/g)  N = 281 | High-Abnormal (>300 mg/g)  N = 49 | p-value |
| --- | --- | --- | --- | --- |
| Primary care, completed (%) | 508 (93.7) | 273 (97.2) | 47 (95.9) | 0.099 |
| Nephrology, completed (%) | 10 (1.8) | 16 (5.7) | 8 (16.3) | <0.001 |
| E-consult with nephrology, Completed (%) | 0 (0.0) | 3 (1.1) | 0 (0.0) | 0.042 |
| ACEi or ARB, new Rx (%) | 22 (4.1) | 19 (6.8) | 2 (4.1) | 0.227 |
| SGLT2, new Rx (%) | 2 (0.4) | 4 (1.4) | 1 (2.0) | 0.167 |
| MRA, new Rx (%) | 8 (1.5) | 4 (1.4) | 0 (0.0) | 0.695 |
| Finerenone, new Rx (%) | 0 (0.0) | 0 (0.0) | 0 (0.0) | NA |
| CKD or proteinuria ICD*, new diagnosis (%) | 7 (1.3) | 40 (14.2) | 10 (20.4) | <0.001 |
| Proteinuria ICD, new diagnosis (%) | 0 (0.0) | 35 (12.5) | 10 (20.4) | <0.001 |
| CKD ICD, new diagnosis (%) | 7 (1.3) | 6 (2.1) | 1 (2.0) | 0.639 |

Supplemental Table 2: Secondary outcomes among those in the diabetes subgroup who completed Minuteful Kidney home ACR test stratified by test result at 1 year from initiation of the study

|  | Normal  (<30 mg/g)  N = 376 | Abnormal (30-300 mg/g)  N = 235 | High-Abnormal (>300 mg/g)  N = 71 | p-value |
| --- | --- | --- | --- | --- |
| Primary care, completed (%) | 345 (91.8) | 216 (91.9) | 70 (98.6) | 0.121 |
| Nephrology, completed (%) | 6 (1.6) | 7 (3.0) | 15 (21.1) | <0.001 |
| E-consult with nephrology, Completed (%) | 0 (0.0) | 0 (0.0) | 1 (1.4) | 0.013 |
| ACEi or ARB, Rx new (%) | 18 (4.8) | 18 (7.7) | 9 (12.7) | 0.035 |
| SGLT2, Rx new (%) | 23 (6.1) | 14 (6.0) | 7 (9.9) | 0.465 |
| mineralocorticoid, Rx new (%) | 2 (0.5) | 0 (0.0) | 2 (2.8) | 0.024 |
| Finerenone, Rx new (%) | 0 (0.0) | 0 (0.0) | 0 (0.0) | NA |
| CKD or proteinuria ICD*, Dx new (%) | 7 (1.9) | 27 (11.5) | 18 (25.4) | <0.001 |
| Proteinuria ICD, Dx new (%) | 0 (0.0) | 19 (8.1) | 14 (19.7) | <0.001 |
| CKD ICD, Dx new (%) | 7 (1.9) | 8 (3.4) | 5 (7.0) | 0.052 |

Supplemental Table 3: Survey questions and responses

| Survey Question |  | N | % |
| --- | --- | --- | --- |
| How easy was it to complete the Minuteful Kidney Test? | Not easy at all | 2 | 0.2 |
|  | Not so easy | 3 | 0.3 |
|  | Somewhat easy | 47 | 5.3 |
|  | Easy | 163 | 18.3 |
|  | Very easy | 675 | 75.8 |
| Are you planning to discuss your test results with your doctor? | I will not share the test results with my doctor | 7 | 0.8 |
|  | Yes during my upcoming visit | 588 | 66.1 |
|  | Yes, I will schedule a visit | 275 | 30.9 |
|  | Other | 20 | 2.2 |
| Did anyone help you complete the test? | I completed the test on my own | 750 | 84.3 |
|  | My formal caregiver | 12 | 1.3 |
|  | My friend or informal caregiver | 15 | 1.7 |
|  | My relative | 113 | 12.7 |
| Did you experience any issues with the kit? | No | 848 | 95.3 |
|  | Yes | 42 | 4.7 |
| How likely is it that you would recommend the Minuteful Kidney test to a friend or colleague? | Not at all likely 0 | 11 | 1.4 |
|  | 1-5 | 89 | 11.0 |
|  | 6-9 | 199 | 24.6 |
|  | Extremely likely 10 | 509 | 63.0 |
| Whose phone did you use to complete the test? | My own phone | 811 | 91.1 |
|  | My formal caregiver’s | 2 | 0.2 |
|  | My friend or informal caregiver’s | 10 | 1.1 |
|  | My relative’s | 67 | 7.5 |
| Would you rather take your urine test at home or at the doctor’s off or lab? | Home | 629 | 70.7 |
|  | Doctor’s office | 61 | 6.9 |
|  | Lab | 28 | 3.1 |
|  | No preference | 172 | 19.3 |
